# Survival differences and artemisinin resistance in severe malaria among HIV coinfected patients: data from Mozambique

**DOI:** 10.64898/2026.06.19.26356090

**Authors:** Irina Mendes de Sousa, Erin McArthur, Lúcia Chambal, Kevan Akrami, Annie N. Cowell, Titos Buene, Robert T. Schooley, Bharti Ajay, Constance A. Benson, Ana Paula Arez, Marcelo U. Ferreira, Elizabeth A. Winzeler, Emília V. Noormahomed

## Abstract

**Background:** Malaria remains a significant cause of morbidity and mortality, especially in sub-Saharan Africa, where rates of HIV coinfection are high. This study aimed to determine whether *Plasmodium falciparum* malaria treatment outcomes and rates of antimalarial resistance markers differ according to HIV serostatus in Mozambique.

**Methodology:** We conducted an observational study of non-pregnant adults, with and without HIV coinfection, admitted to the Hospital Central de Maputo for treatment of severe malaria. *Plasmodium falciparum* DNA was extracted from whole blood and sequenced to identify single-nucleotide polymorphisms. Statistical analyses to compare clinical outcomes and rates of nonsynonymous mutations in genes associated with drug resistance were performed in R version 4.2.

**Results:** We recruited 149 study participants aged between 18-62 years, 72 (48.3%) were female, and 59 (39.6%) were infected with HIV. Comparing clinical outcomes, we found a significant difference in anemia (hemoglobin <5 mg/dl: P-value = 0.01248) but no other predefined clinical characteristics including fever, renal dysfunction, or pulmonary edema in those with and without HIV coinfection. We found significantly higher in hospital mortality in those with HIV coinfection (*P*-value = 0.03416, and at 30 days post discharge *P*-value = 0.007526). Mortality correlated inversely with CD4 count. Overall mortality at the end of the study was 20.1%. We did not find a significant increase in the number of nonsynonymous mutations in known antimalarial resistance associated genes *pfmdr1*, *pfcrt*, *pfdhfr*, *pfdhps*, in patients who reported taking antimalarials prior to admission or in those with HIV coinfection. However, we did find an increase in rates of K189T *pfkelch* mutations (21.2% vs 2.7%, p = 0.022) in HIV infected patients. We found no increase in rates of nonsynonymous mutations in *pfdhfr* in HIV infected patients taking TMP-SMX prophylaxis: *pfdhfr-pfdhps* quintuple mutations were found in almost 100% of the parasite population.

**Conclusions:** We find a higher mortality rate in patients with malaria-HIV coinfection, further emphasizing the importance of effective treatment for this vulnerable population. We also provide an important baseline for the rate of resistance-conferring mutations in a population at risk of harboring resistant parasites in Mozambique, highlighting the need for continued surveillance.

## Introduction

HIV-malaria coinfection exacerbates the burden and severity of both diseases and increases the complexity of clinical management (1). The geographic distribution of these diseases overlaps particularly in sub-Saharan Africa (SSA) (2). The burden of HIV-malaria coinfection has particularly dire public health consequences in Mozambique, where both diseases are exceedingly prevalent. Data collected by the National Monitoring and Evaluation System (SIS-MA) report that in 2024, 11.6 million cases of malaria were reported in outpatient consultations and 66.8 thousand cases were admitted for hospitalization, while 358 in-hospital deaths were reported nationwide. This small country alone accounts for 4% of the current global malaria burden (3).

In addition, Mozambique has one of the greatest prevalence rates of HIV worldwide, estimated at 12.4% among people aged 15-45 years, corresponding to approximately 2,097,000 HIV-infected adults countrywide, according to the Mozambique Population-Based HIV Impact Assessment 2021 (4). HIV prevalence reaches 15.4% in the Province of Maputo, where the country’s capital and largest city is located. Approximately 63,000 new annual HIV infections are estimated to occur among adults (4).

Malarial infection in Mozambicans living with HIV ranges from subclinical to life-threatening disease (5). HIV coinfection has been associated with increased malaria severity and mortality, particularly in children and pregnant women (6–8). Published literature reporting the severity of clinical outcomes in coinfected nonpregnant adults is less consistent.

Although some studies have reported no differences in outcomes between adults with and without HIV with severe malaria (9), many others, particularly in SSA, report HIV coinfection to be correlated with worse outcomes in pregnant as well as nonpregnant adults (2,7). One prior hospital-based study carried out in Maputo reported higher parasite densities and increased frequencies of respiratory distress, abnormal bleeding, jaundice, renal impairment, and hypoglycemia, with a greater in-hospital mortality due to *P. falciparum* malaria (13% vs. 2%), in HIV-coinfected compared with HIV-uninfected adult patients (8).

Malaria and HIV interact to cause dysregulation of the immune system. Independently, HIV and malaria interact with the host immune system to trigger complex activation of immune cells, which causes altered cytokine levels and antibody production (2,10). Furthermore, HIV targets CD4^+^ T cells, which play an important role in the development and maintenance of antimalarial immunity. Mechanisms of dysregulation include widespread lymphoid necrosis, hyperactivation of CD4^+^ and CD8^+^ T cells to secrete cytokines, and HIV-induced reduction in CD4^+^ T cells. In summary, HIV and malaria coinfection work together to enhance immune dysregulation both via suppression and hyperactivation, leading to increased severity of presentation of both infections (2,11).

In addition to the described effect of HIV infection on increasing the severity of *Plasmodium* spp. infection, HIV, and antiviral therapies also alter antimalarial drug metabolism, impairing the protection offered by antimalarial treatment and increasing rates of adverse reactions. Furthermore, this supports the selection of mutant strains associated with resistance to antimalarial drugs, facilitating increased burden of disease and rates of malaria transmission (2,10,12).

Antimalarial resistance constitutes one of the biggest barriers to controlling malaria in endemic countries (13). Interactions between several antimalarials and antiretroviral drugs have been reported to affect the efficacy of artemisinin-based combination therapies (2,14–16). For example, Efavirenz (EFV), a non-nucleoside reverse transcriptase inhibitor (NNRTI), is a known CYP450 inducer leading to increased metabolism of both artemether and lumefantrine (16,17). Similarly, Nevirapine (NVP) co-administration with artemether-lumefantrine leads to decreased and subtherapeutic artemether levels (18,19). This has also been described for protease inhibitors. Co-administration of Lopinavir (LPV) and artemether-lumefantrine results in decreased artemether but increased lumefantrine blood concentrations (20,21).

Accumulating evidence from the past decades mostly refers to the effect of HIV on malaria in the absence of effective antiretroviral therapy (ART). However, as of 2022, 91% of all adults with HIV from low and middle-income countries have been receiving dolutegravir-based ART regimens recommended by the World Health Organization (22). This represents a major shift from earlier treatment policies, as in 2016 the first-line ART regimen in Mozambique was still predominantly efavirenz based (23). Data collection in our study found 61% of participants with HIV were on ART (Table 1), aligning with data from other studies in this period. In Mozambique in 2021, 69% of the adults living with HIV were on ART and 96% of those were aware of their HIV-positive status according to the Instituto Nacional de Saúde (INS) in Mozambique (4).

**Table 1.** Coinfection rates according to demographic, clinical features and antiretroviral regimen of study participants.

|  | HIV infected<br>(Total = 59) | HIV non-infected<br>(Total = 82) | *P*-value<sup>a</sup> |
| --- | --- | --- | --- |
| **Characteristic** |  |  |  |
| Age group* [No. (% of total)] |  |  |  |
| 18-40 years | 32 (54.2) | 42 (51.2) | 0.923 |
| 41-59 years | 21 (35.6) | 21 (25.6) |  |
| 60+ years | 6 (10.2) | 16 (19.5) |  |
| Female sex [No. (% of total)] | 29 (49.1) | 41 (50.0) |  |
| **No. (%) on antiretroviral therapy (ART) on admission** |  |  |  |
| TDF+3TC+EFV | 29 (80.6) |  | 0.030 |
| AZT+3TC+NVP | 4 (11.1) |  |  |
| ABC+3TC+EFV | 2 (5.6) |  |  |
| AZT + 3TC + Lopinavir/R | 1 (2.8) |  |  |
| **Other medication history** |  |  |  |
| Antimalarial use prior to admission, n (%) | 21 (35.6) | 15 (18.3) | 0.033<sup>b</sup> |
| Cotrimoxazole on admission, n (%) | 25 (42.4) | 1 (1.2) | 2.50e-08<sup>c,d</sup> |
| **CD4 cell count (cells/μL)<sup>b</sup>** |  |  |  |
| No. (%) <350 cells/μL | 41 (69.5%) | | |
| **Parasite density (%)<sup>e</sup>** |  |  |  |
| % Parasitemia | 0.70 (0.2-5.68)% | 1.60 (0.2-15.56)% | 0.067<sup>f</sup> |
| **Blood cell counts (cells/μL)<sup>#</sup>** |  |  |  |
| White blood cells (WBC) | 7956 (600-23000)/μL | 7491.5 (100-37300)/μL | 0.591<sup>f</sup> |
| Platelets | 73084.7458 (4000-346000)/μL | 87314 (4,000-516,000)/ μL | 0.267<sup>f, g</sup> |
| Hemoglobin, g/100 mL | 7.8 (2.8-14.9) g/100 mL | 9.5 (3.6-16.3) g/100 mL | 0.005<sup>f</sup> |

Demographic and clinical data for 149 subjects admitted to Maputo Central Hospital with severe malaria. Overall, 59 (39.6%) patients were confirmed to have HIV coinfection and 8 patients had an undetermined HIV status.
\*Missing data from 2 subjects
<sup>#</sup>[mean (range)]
<sup>a</sup> *P* value for chi-square test for a comparison between HIV infected and HIV non-infected patients unless otherwise specified
<sup>b</sup> Data from 58 HIV infected and 82 HIV non-infected participants
<sup>c</sup> *P* value for Fisher's exact test for a comparison between HIV infected and HIV non infected patients
<sup>d</sup> Data from 54 HIV infected and 68 HIV non-infected participants
<sup>e</sup> Data from 45 HIV infected participants; measurements were done at HCM (37 patients) or other health facilities (8 patients).
<sup>f</sup> *P* value for Student's *t* test for a comparison between HIV infected and HIV non-infected patients

Whether HIV-coinfection continues to pose a substantial risk of malaria complications and mortality in sub-Saharan Africa in the current era of widely available and affordable ART is uncertain. Here, we address this knowledge gap by comparing the clinical features and prevalence of adverse outcomes in a series of HIV-infected and HIV-uninfected adults with severe malaria admitted to the largest hospital in Mozambique. We also compare antimalarial therapy related gene mutations across population subsets. From the results, we hope to inform recommendations regarding the management of malaria in the context of HIV.

## Methods

### Study site and design

This study was conducted at the Hospital Central de Maputo (HCM), a 1,200-bed quaternary teaching hospital of the Faculty of Medicine of Eduardo Mondlane University in Maputo, Mozambique (24). Maputo city (population, 1,136,296) is located in the southern part of the country, on the west side of Maputo Bay. HCM serves as a referral center for residents in Maputo city and patients from all over the country (24). Mozambique has two main seasons: a hot and rainy season (November to March) and a dry season (April to October). Despite these seasonal fluctuations, the tropical climate in Mozambique presents patterns of precipitation, temperature, and humidity that lead to the continuous transmission of malaria throughout the year (25).

This single-center cross-sectional study included non-pregnant adult patients >18 years of age admitted to MCH with severe malaria (defined below). Between December 2016 and Marchm 2018, we enrolled 149 patients who presented to the Emergency Department with *P. falciparum* infection confirmed by a First Response® Malaria Antigen *P. falciparum* (HRP2 - 91% sensitivity) or thick-blood smear microscopy (100 microscopic fields). Based on clinical assessment, patients were admitted to either the intensive care unit (ICU) or medicine wards. This study was approved by the National Bioethics Committee of Mozambique and the Human Research Protections Program of the University of California, San Diego (UCSD).

### Laboratory measurements

Malarial infection was confirmed by expert microscopy. Two technicians independently read Giemsa-stained thick smears prepared on admission, and a third microscopist, blinded to the original results, examined any slides with discordant results. Parasitemia was estimated as follows: parasites/μL = parasite count per 200 white blood cells (WBC) × WBC count/μL measured on admission/200 (26). Self-reported HIV infection status was confirmed by laboratory testing upon admission (see below). CD4^+^ T-lymphocyte counts were determined only for HIV infected participants. Samples of whole blood were frozen for shipment to UCSD, IGM Genomic Center for further processing.

### Case definitions

Severe malaria was defined by the presence of one or more modified World Health Organization clinical and laboratory criteria (27) among patients with laboratory-confirmed *P. falciparum* infection and no other identified alternative cause. Severity criteria included (a) hyperparasitemia [two tentative parasite density thresholds: >100,000 *P. falciparum* blood stages/μL (approximately 2.5% parasitemia) and >200,000/μL (5% parasitemia)], (b) unrousable coma (Glasgow coma score < 9), (c) multiple convulsions (>2 episodes within 24 h), (d) metabolic acidosis (plasma bicarbonate <15 mmol/L), (e) severe anemia (hemoglobin <5 g/100 mL), (f) evidence of renal impairment (plasma creatinine >265 µmol/L and/or urine output <400 mL within 24 h), (g) hypotension (systolic blood pressure <70 mmHg); (h) radiologically confirmed pulmonary edema, (i) jaundice (total plasma bilirubin >50 µmol/L), and (j) abnormal bleeding, including recurrent or prolonged bleeding from the nose, gums, venipuncture sites, or hematemesis or melena.

HIV positivity was defined as a previously documented HIV positivity or a positive **Alere™ HIV 1/2 test** (Alere Medical Co., USA), a rapid immunochromatographic assay, followed by a positive **Uni-Gold™ HIV** test (Trinity Biotech, Ireland), on admission. Most HIV-infected patients had CD4^+^ lymphocyte counts determined on admission at HCM using flow cytometry; otherwise, we considered the most recent available CD4^+^ count. Viral load estimates were not available.

### Generation of Whole Genome Sequences data and variant calling

Samples from 120 subjects were shipped on dry ice from Maputo to UCSD. All methods regarding selective whole genome amplification (SWGA) and variant calling have been described in detail in an earlier publication (28). Variants were annotated using SnpEff, with focus on missense mutations in genes previously associated with drug resistance (*pfcrt, pfmdr1, pfdhfr, pfdhps, pfkelch13,* and *pfaat1*).

### Data Analysis

Data were collected using REDCap (29) and analyzed using R version 4.2 (R Core Team (2022). *R: A language and environment for statistical computing*. R Foundation for Statistical Computing. https://www.R-project.org/). For comparing clinical outcomes with dichotomous classifications, P-values are from chi squared tests, or Fisher’s exact test (when count <5 in any group). When comparing missense mutations in known drug resistance genes across population subsets including differing HIV status or prior drug exposure, P-values are from Fisher’s exact tests.

## Results

### Demographic and Clinical Characteristics

Demographic and clinical data were available for 149 subjects (Table 1). Subjects had an average age of 42.3 years (18–62): 48.3% were women and 51.7% men. Of this cohort, 40.9% had known HIV infection, and 5.4% had unknown HIV infection status due to missing confirmatory testing.

In the HIV infected group, thirty-six participants (61.0%) were taking antiretroviral therapy (ART) on admission. ART regimens included tenofovir (TDF)-lamivudine (3TC)-efavirenz (EFV), zidovudine (AZT)-3TC-nevirapine (NVP), abacavir (ABC)-3TC-EFV, and AZT-3TC-lopinavir-ritonavir (LPV/r) (Table 1). Only 25 HIV-infected patients (42.4%) were reportedly taking co-trimoxazole when admitted to HCM. Although nearly two-thirds of the HIV infected patients were on ART, the mean CD4^+^ cell count was 178/μL (95% CI, 119-176/μL). Only 8.8% of the 45 HIV-infected individuals with recent measurements had more than 350 CD4^+^ cells/μL (Table 1).

On admission, all but two patients (98.6%) received intravenous artesunate at the HCM Emergency Room (standard dose of 2.4 mg/kg of body weight). One (HIV non-infected) patient was given intravenous quinine followed by oral quinine-doxycycline and one (HIV infected) was initially given oral artemether-lumefantrine followed by intravenous artesunate upon transfer to the medicine ward. Notably, no patients had persistent parasitemia after 72 hours of hospitalization.

Overall mortality was 20.1% including in-hospital mortality and within 30 days of discharge.

### HIV status vs clinical outcome

Clinical indicators and outcomes between study participants with severe malaria with or without HIV were compared. We found no significant difference in the severity of predetermined clinical symptoms such as of pulmonary edema, mental status (CGS score <9), evidence of abnormal bleeding or disseminated intravascular coagulations (DIC), hypotension (SBP<70 mmHg), fever greater than 40 °C, elevated blood urea nitrogen (BUN>15 mg/dL) or bilirubin (>2.5 mg/dl) between populations (Table 2). However, patients with HIV coinfection were significantly more likely to have a hemoglobin <5 mg/dL compared to those without HIV (P-value = 0.01248, via Pearson’s chi-squared test).

**Table 2.** Clinical features and outcomes of study participants.

| **Clinical symptoms on admission** | **HIV infected (n = 59)** | **HIV non-infected (n = 82)** | **P-value<sup>a</sup>** |
| --- | --- | --- | --- |
| Hypotension (SBP<70 mmHg) | 1 (1.69) | 3 (3.66) | 0.6335<sup>b</sup> |
| Fever (T >=40°C) | 17 (28.8) | 22 (26.6) | 0.9613 |
| Abnormal bleeding or DIC | 3 (5.1) | 1 (1.22) | 0.3158<sup>b</sup> |
| Total Bilirubin > 2.5 mg/dl | 12 (20.3) | 14 (17.1) | 0.7418 |
| GCS <9 | 18 (30.5) | 14 (17.1) | 0.09168 |
| Urea >15 mg/dL | 31 (52.6) | 30 (36.6) | 0.1137 |
| Hgb <5 mg/dL | 20 (33.9) | 12 (14.6) | 0.01248 |
| Pulmonary edema | 5 (8.5) | 1 (1.22) | 0.0849<sup>b</sup> |
| **Clinical outcomes** |  |  |  |
| Survival on discharge | 48 (81.4) | 73 (89.0) | 0.03416 |
| Survival at 30d<sup>c</sup> | 40 (67.8) | 66 (80.5) | 0.007526 |

HIV infected patients were significantly more likely to have a Hgb <5 mg/dL, and had higher rates of mortality prior to discharge and at 30d post discharge. P-values are from chi squared tests, or fisher's exact test (when count <5 in any group).
<sup>a</sup> P-value for a chi-square test for a comparison between HIV infected and HIV non-infected patients unless otherwise specified
<sup>b</sup> P-values calculated via Fisher's exact test
<sup>c</sup>Data from 56 HIV infected and 79 HIV non-infected patients

Regarding mortality, patients with previously documented HIV infection or with positive rapid test on admission had significantly higher levels of mortality prior to discharge (P-value = 0.03416, Pearson’s chi-squared test) and at 30 days post discharge (P-value = 0.007526, Pearson’s chi-squared test) (Table 2).

When comparing patient CD4 count on admission to survival, we found a significant decrease in survival in those with CD4 counts less than 100 cells/μL. Those with CD4 counts <100 cells/μL had a 30-day probability of survival <35%, while those with CD4 counts >150 cells/μL had 30-day probability of survival >80%, similar to participants without HIV coinfection (Figure 1).

**Figure 1:**
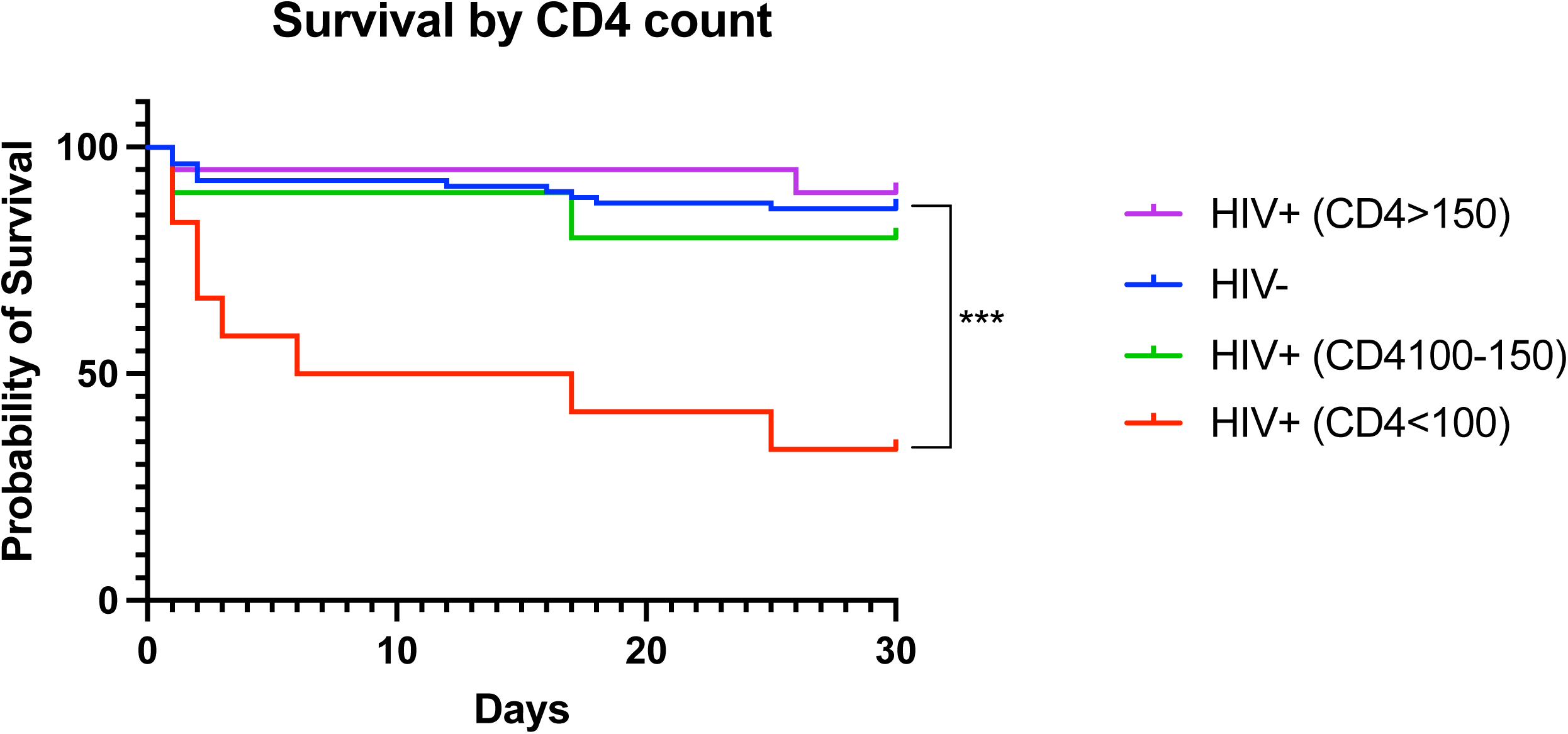
Survival by CD4 count. There was no statistical difference in survival between HIV non-infected patients and those with CD4 >150 cells/μL (p = 0.6632) or CD4 count between 100-150 cells/μL (p = 0.5779). However, there was a statistical difference in survival between HIV non-infected patients and those with CD4 <100 cells/μL (p<0.0001). Statistical analysis performed via Log-rank (Mantel-Cox) test.

### Artemisinin resistance evaluation Antimalarial pretreatment

We compared rates of mutations in resistance conferring genes between patients who reported taking antimalarials prior to admission and those who did not. Of patients from whom sequences were obtained, 26 reported taking antimalarials prior to admission. Most often previous treatment was with artemether lumefantrine (Coartem) (20), followed by artesunate (4), artesunate + doxycyline (1), and Coarsucam (ASAQ) (1). We did not find any significant increase in the rates of missense mutations in *pfmdr1, pfcrt, pfdhfr, pfdhps, pfkelch13* or *pfaat1* in patients who reported taking antimalarials prior to admission (Supplementary Table 1).

### HIV status vs molecular markers of resistance

HIV infected patients were more likely to be taking Trimethoprim/sulfamethoxazole (Co-trimoxazole) on admission (P-value = 4.427e-9) which was hypothesized to result in increased mutation rate in genes conferring antifolate resistance, such as the dihydrofolate reductase (*pfdhfr*) gene. However, we find no difference in the rates of nonsynonymous mutations in *pfdhfr* between HIV-infected and noninfected patients, owing to the near 100% prevalence of *pfdhfr-pfdhps* quintuple mutations in the parasite population HIV infected patients were also more likely to have taken antimalarial medications prior to admission (P-value = 0.02948); however, we did not find a statistically significant increase in missense mutations in *pfcrt*, *pfdhps*, *pfmdr1,* or *pfkelch13*. (Table 3). Rates of K189T mutation in *pfkelch13* in the HIV infected group were notably higher, 21.2% compared with 2.7% in the HIV non-infected group, with a significant p-value (p = 0.022). Although associated with delayed parasite clearance in population studies in Nigeria(30), this mutation is not in the kelch13 protein propeller domain, the site of most mutations that have been confirmed in vitro to play a role in artemisinin resistance(31).

**Table 3:** Prevalence of missense mutations in drug resistance genes among study participants.

| Proteins name | Description | HIV infected AA substitution (#variants/total callable) | HIV non-infected AA substitution (#variants/total callable) | P value<sup>a,b</sup> |
| --- | --- | --- | --- | --- |
| DHFR-TS | Bifunctional dihydrofolate reductase-thymidylate synthase | p.N51I (33/33)<br>p.C59R (33/33)<br>p.S108N (32/32) | p.N51I (39/39)<br>p.C59R (38/39)<br>p.S108N (39/39) | NS<br>NS<br>NS |
| PPPK-DHPS | Hydroxymethyldihydropterin pyrophosphokinase-dihydropteroate synthase | p.S436A/C(1/35)<sup>a</sup><br>p.G437A (2/35)<br>p.K540E (31/34) | p.S436A/C(1/38)<sup>b</sup><br>p.G437A (2/38)<br>p.K540E (35/38) | NS<br>NS<br>NS |
| MDR1 | Multidrug resistance protein 1 | p.Y184F (15/33)<br>p.D642G (3/32)<br>p.N652D (13/30) | p.Y184F (17/36)<br>p.D642G (2/34)<br>p.N652D (18/34) | NS<br>0.67<br>0.46 |
| CRT | Chloroquine resistance transporter | p. D24Y (1/34) | p. D24Y (0/38) | NS |
| KELCH13 | Kelch protein K13 | p. K189T (7/33)<br>p. T192P (1/34)<br>p. R255K (0/34) | p. K189T (1/37)<br>p. T192P (0/38)<br>p. R255K (1/39) | 0.022<br>0.47<br>NS |

Among the mutations analyzed, only the K189T variant in *pfkelch13* showed a significant difference in prevalence between the HIV infected and HIV non-infected groups.
<sup>a</sup>P values calculated using Fischer exact test
<sup>b</sup>NS = $p > 0.9$
<sup>c</sup>Sensitive genotype: reference strain 3d7 is sulfadoxine resistant with an A437G mutation

## Discussion

In this study, we investigated clinical outcomes and artemisinin resistance patterns in patients co-infected with HIV and severe malaria in Maputo, Mozambique. Our results show an overall mortality rate of 20.1%, including in-hospital mortality and within 30 days of discharge, with a significant difference in survival between HIV infected and non-infected patients. These results highlight the need for an increased level of care and prompt effective treatment for malaria patients co-infected with HIV, especially in those with CD4 counts <100 cells/μL.

Our study found no significant increase in mutation rates in malaria resistance-related genes for patients who reported antimalarial use prior to admission. Our results suggest that although HIV infection may influence the severity and increase the risk of mortality in patients with severe malaria, we found no evidence that it is directly associated with antimalarial drug resistance. However, additional studies are needed to fully understand the impact of coinfection on antimalarial drug resistance. We also suspect that many SNP rates already reaching fixation in the parasite population obscures differences we might have found between HIV serostatus groups.

### HIV status vs clinical outcome

In general, in clinical terms, those with HIV had higher proportions of severity-defining parameters used in the study: pulmonary edema, mental status (CGS score <9), evidence of abnormal bleeding or disseminated intravascular coagulations (DIC), elevated blood urea nitrogen (BUN>15 mg/dL) or bilirubin (2.5 mg/dL) and anemia (hemoglobin<5 mg/dL). Similar findings have been reported by other studies in Mozambique (6,8) and neighboring countries such as South Africa (15,32) and Zambia (33). However, none of these differences reached statistical significance in our study, other than anemia, which is a known sequela of HIV infection (34). However, this degree of anemia suggests that hemoglobin levels are also likely exacerbated by the severity of malaria illness perpetuated by HIV infection, rather than HIV infection alone, and anemia is known to be significantly associated with malaria severity (6,9,35).

Previous studies have reported HIV status to modulate the severity of sequelae of malaria infection (8,32). However, in our study, enrolled patients already met criteria for severe malaria and per definition were exhibiting clinical symptoms of severe disease. This likely resulted in masking the ability of HIV serostatus to result a further increase in severity of symptoms. It is also possible that our sample size was not large enough to reach statistical significance given this effect of studying severe malaria.

We observed a significant difference in mortality rates between those with and without HIV infection. Our study corroborates what others have shown that HIV infection confers an increased risk of mortality in patients with severe malaria (36). Furthermore, we show that patients with advanced HIV and CD4 counts less than 100 cells/μL had significantly higher rates of mortality than those with CD4 counts >150 cells/μL. It is important to consider possible factors that may contribute to the disparities in outcomes observed between patients co-infected with HIV and malaria. Immunosuppression caused by HIV can compromise the ability of the host’s immune system to control the replication of the malaria parasite, leading to an exacerbated inflammatory response and greater severity of the disease (2,10). Also, the presence of other comorbidities associated with HIV, such as opportunistic infections, which occur at higher frequency with lower CD4 counts can complicate or influence clinical outcomes of patients (37). Furthermore, severe anemia associated with HIV infection can contribute to morbidity and mortality in patients with severe malaria, increasing the risk of complications (8,34). HIV co-infection may also have implications for the clinical management of patients with severe malaria, as patients with HIV may have a suboptimal response to antimalarial treatments due to altered pharmacokinetics or drug interactions with antiretroviral therapies (2,14–16).

However, factors such as the immunological status of the patients, adherence to antiretroviral treatment, and promptness in diagnosing and treating malaria can significantly influence clinical outcomes (7). Integrated health care strategies that address both HIV infection and malaria, such as integrating screening and treatment services, may be essential to improve outcomes in co-infected patients.

### Antimalarial resistance evaluation

When investigating antimalarial resistance, we compared the rates of mutations in resistance-related genes between patients who reported using antimalarials before admission and those who did not. We hypothesized that severe malaria patients reporting recent treatment may have a higher likelihood of carrying resistant parasites. However, we did not find any significant differences in rates of resistance conferring mutations in patients reporting prior antimalarial use. This could be due to our study limitations including challenges in reporting, small sample size, and our screening a limited number of genes. We found a significant difference in rates of antimalarial pretreatment in patients with HIV coinfection. This could likely be attributed to increased contact with healthcare system and access to treatments. It could also indicate higher rates of treatment failure in HIV infected patients due to the mechanisms previously described such as drug-drug interactions between antimalarials and antiretrovirals resulting in inadequate plasma concentrations of antimalarial medications.

We also hypothesized that patients with HIV coinfection might be at increased risk of harboring resistant parasites due to reduced immune pressure allowing less-fit, drug-resistant parasites to survive and replicate, as well as recurrent or prolonged infection, and exposure to cotrimoxazole prophylaxis (38,39). However, we found no significant differences comparing rates of known resistance conferring mutations between HIV infected and non-infected populations.

We did find a significant, 10-fold increase in rates of *pfkelch* K189T mutations in the HIV infected population compared to the HIV non-infected group. While N-terminal K189T mutations have been associated with delayed parasite clearance(40), to our knowledge, there are no reports in which CRISPR-Cas9 genome editing has been used to demonstrate that an isolated K189T allele confers resistance to the endoperoxide class (artemether, artesunate, OZ439, DHA, etc.) of antimalarial in a laboratory setting. In addition, the mutation is not located in the Kelch13 protein’s propeller domain (amino acids 430–723), the domain associated with most PfKelch13 mutations that have been proven to confer resistance in a laboratory setting(31). Nevertheless, a recent study showed that laboratory-provided artemisinin drug pressure could select for a P413A mutation in the non-propeller BTZ/POZ domain of *pfkelch13*(41) . Furthermore, most studies on pfkelch13 involve testing with dihydroartemisinin (DHA), and in Mozambique artemether is used. Finally, the standard Ring-Stage-Survival assay(42) that is used to study propeller domain mutations may not necessarily reveal phenotypic changes from non-propeller mutations. Thus, more work is needed.

If indeed the K189T mutation has a real impact, its overrepresentation in HIV populations could be due to the several reasons. We hypothesize that patients living with HIV could be at higher risk of harboring resistance parasites due to decreased immune pressure, allowing for propagation of resistance mutations that carry a fitness cost such as K189T. As such, this population could provide a unique reservoir of parasites under decreased survival pressure from the host immune system. These patients should continue to be evaluated closely for emerging drug resistance and the development of de novo mutations.

It is also possible that increased exposure to the medical interventions in these patients could lead to increased exposure to prior antimalarial treatment. While there was no notable difference in mutation rates between patients who had recently taken antimalarial prior to admission, we do not know about previous history of antimalarial treatment for these patients. We know that Cotrimoxazole exposure was higher in the HIV positive population and that cotrimoxazole is associated with mutations in *pfdhfr* and *pfdhps,* that due to its disruption of the folate pathway. However, we are not aware of any biological mechanism by which the K189T mutation would confer a selective advantage under cotrimoxazole pressure. ART exposure itself may impose selective pressure, as protease inhibitors and NNRTIs have documented antiparasitic effects. This could favor parasites harboring polymorphisms such as K189T, or alternatively, K189T may be linked to other mutations under selection by ART exposure.

Finally, it is possible that differences in malaria transmission intensity between populations of HIV infected and HIV non-infected individuals can create confounding effects that influence the observed prevalence of certain parasite mutations, including K189T, related to delayed parasite clearance (30). Lower malaria transmission settings often overlap with areas of higher HIV burden.

In the case of *pfdhps* and *pfdhfr,* a lack of difference in mutation rates was due to rates reaching fixation across both populations. We hypothesized that higher rates of cotrimoxazole exposure in the HIV infected group would result in higher rates of sulfoxide resistance (43). In our data, the homogeneously resistant parasite population inhibited our ability to differentiate mutation rates between populations.

## Conclusion

In conclusion, our data contribute to the growing body of data revealing increased mortality risk in malaria patients with HIV coinfection. In addition to increased risk of death, these patients have many risk factors for the development of parasite drug resistance including prolonged infection due to immunosuppression, exposure to prophylaxis with sulfonamides, and drug interactions resulting in suboptimal antimalarial dosing. Despite these risk factors, our analysis reports no significant increase in known drug resistance conferring SNPs in HIV coinfected patients. As mentioned, this could be due to limitations of the study such as a small sample size and the differences noted in rates of K189T mutations warrant further investigation. Additionally, our screen was limited to known resistance conferring SNPs and it is possible that these patients harbor yet undiscovered mutations affecting treatment efficacy which would also provide an additional explanation for the worse outcomes observed in coinfected patients receiving treatment for severe malaria. Overall, this work highlights the importance of effective treatment and close surveillance in this high-risk population.

## List of abbreviations

SSA: Sub-Saharan Africa
EFV: Efavirenz
NNRTI: Non-nucleoside reverse transcriptase inhibitor
NVP: Nevirapine
LPV: Lopinavir
ART: Antirretroviral therapy
INS: Instituto Nacional de Saúde amplification

## Declarations

### Ethics approval and consent to participate

This study was approved by the National Bioethics Committee of Mozambique and the Human Research Protections Program of the University of California, San Diego (UCSD) and followed the principles of the Helsinki Declaration. Prior to the initiation of the study, participants over 18 years of age were explained about the study aims and methodology in order to obtain their written informed consent. The written informed consent was also obtained from the legal guardian of each participant unable to consent. Illiterate participants were asked to add their fingerprint to the consent form along with an impartial witness who also signed and was present during the informed consent process. All information of the participants were kept strictly confidential.

### Consent for publication

All authors read and approved the final version of the manuscript.

### Availability of data and materials

The datasets used and/or analyzed during the current study are available from the corresponding author.

## Authors’ contributions

Conceptualization: IMS and EM, ANC, RTS, CAB, EW, EVN; Data curation: IMS, EM, and MF; Funding acquisition: EW, RTS and EVN; Methodology: IMS and EM, LC, ANC, TB, RTS, CAB, EW, EVN; Project administration: EW, RTS and EVN. Supervision: EW, RTS, APA and EVN; Writing original draft: IMS and EM, LC, KA, ANC, TB, RTS, BA, CAB, APA, MUF, EW and EVN; Review and editing: IMS and EM, ANC, RTS, CAB, APA, MUF, EW and EVN.

## Data Availability

All data produced in the present study are available upon reasonable request to the authors.

## Acknowledgements

We are indebted to all patients who contributed to this study. We also acknowledge to the staff of Maputo Central Hospital, who helped to identify study participants and supported the collection of data. The authors are also grateful to Professor Virgilio do Rosário, retired Professor at the Instituto of Tropical Medicine and Hygiene from Portugal for the insights provided to develop and conduct the present research.

## Additional Files

Additional File 1: Supplementary Table 1 (.docx):

**Table S1: Prevalence of missense mutations in drug resistance genes, comparing effects of reported antimalarial exposure prior to admission.**

