## Supplemental Table 1 for "Survival differences and artemisinin resistance in severe malaria among HIV coinfected patients: data from Mozambique"

**Table S1:** **Prevalence of missense mutations in drug resistance genes, comparing effects of reported antimalarial exposure prior to admission.**

| **Proteins name** | **Description** | **No antimalarial exposure PTA AA substitution (#variants/total callable)** | **Antimalarials reported PTA AA substitution (#variants/total callable)** | ***P* value^a,b^** |
| --- | --- | --- | --- | --- |
| DHFR-TS | Bifunctional dihydrofolate reductase-thymidylate synthase | p.N51I (57/57)  p.C59R (56/57) p.S108N (57/57) | p.N51I (18/18)  p.C59R (18/18) p.S108N (17/17) | NS  NS  NS |
| PPPK-DHPS | Hydroxymethyldihydropterin pyrophosphokinase-dihydropteroate synthase | p.S436A/C(2/57)  p.G437A (4/56) p.K540E (50/56) | p.S436A/C(1/21)^c^  p.G437A (1/22) p.K540E (20/21) | NS  NS  0.69 |
| MDR1 | Multidrug resistance protein 1 | p.Y184F (23/55)  p.D642G (3/54)  p.N652D (23/54) | p.Y184F (10/18)  p.D642G (2/18)  p.N652D (8/16) | 0.39  0.60  0.83 |
| CRT | Chloroquine resistance transporter | p. D24Y (1/54) | p. D24Y (0/18) | NS |
| KELCH13 | Kelch protein K13 | p. K189T (6/57)  p. T192P (1/58)  p. R255K (1/58) | p. K189T (2/16)  p. T192P (0/17)  p. R255K (0/18) | NS  NS  NS |

Among the mutations analyzed, there was no significant different between mutation rates comparing those with and without reported antimalarial use prior to admission.

^a^P values calculated using Fischer exact test

^b^NS = p > 0.9

^c^Sensitive genotype: reference strain 3d7 is sulfadoxine resistant with an A437G mutation
